# Half the picture: Word frequencies reveal racial differences in clinical documentation, but not their causes

**DOI:** 10.1101/2022.02.10.22270631

**Authors:** Jacqueline A. Penn, Denis Newman-Griffis

## Abstract

Clinical notes are the best record of a provider’s perceptions of their patients, but their use in studying racial bias in clinical documentation has typically been limited to manual evaluation of small datasets. We investigated the use of computational methods to scale these insights to large, heterogeneous clinical text data. We found significant differences in negative emotional tone and language implying social dominance in clinical notes between Black and White patients, but identified multiple contributing factors in addition to potential provider bias, including mis-categorization of some healthcare vocabulary as emotion-related. We further found that notes for Black patients were significantly less likely to mention opioids than for White patients, potentially reflecting both inequitable access to medication and provider bias. Our analysis showed that computational tools have significant potential for studying racial bias in large clinical corpora, and identified key challenges to providing a nuanced analysis of bias in clinical documentation.

## Introduction

Addressing racial disparities in healthcare requires that mechanisms of disparity be identified and continually monitored. Informatics technologies have the potential to be tools for combatting racial disparities in healthcare, but can also serve to amplify racial biases in the health data they analyze. Understanding what health data are saying— and implying—about patients and the care they receive is key both to improving the quality and equitability of that care and to building equitable informatics technologies. While structural factors such as income inequality are major contributors to racial health disparities, implicit racial biases exhibited by healthcare providers also play a significant role due to their effect on clinical decision-making and patient-provider interactions.^1^ Clinical documentation is the best record of providers’ perceptions of their patients, and is central to clinical decision making throughout the course of care. Thus, understanding what implicit bias looks like in clinical documentation is key to both improving provider education, such as modifying medical school curriculums or Continuing Medical Education (CME) for licensed professionals, and designing equitable natural language processing (NLP) methods to analyze the invaluable information in clinical notes.

The impacts of implicit bias on medical care and health outcomes are significant. Previous studies have found that clinicians with higher levels of pro-White implicit bias, as measured by Implicit Association Tests (IATs), are less likely to, relative to White patients, treat Black patients with thrombolytics^2^ or narcotics,^3, 4^ less likely to refer Black patients with chest pain to a specialist^5^, and more likely to diagnose Black patients with less-severe disease.^5^ The pain of Black patients is often underestimated and undertreated, likely due in part to common myths that persist in healthcare. A 2016 study found that many medical students and residents believed myths about Black patients, such as that they have thicker skin or less sensitive nerve endings than White patients. Furthermore, the authors found that belief in such race-based medical myths was correlated with less accurate treatment decisions for Black patients.^6^

Previous studies have shown that provider language can reflect these racial biases. Beach et al.^7^ found higher levels of disbelief-related language in clinical notes for Black and female patients, supporting the idea that Black and female patients’ complaints are not taken as seriously as White or male patients. Hagiwara et al.^8^ analyzed transcriptions of physician-patient interactions, and found greater use of anxiety-related language and first-person plural pronouns (an indicator of social dominance) in racially-discordant interactions by physicians with greater levels of IAT-measured implicit bias. Park et al.^9^ also looked at social dominance by hand-analyzing 600 clinical notes and found social dominance to be one of the ways clinicians express negative emotions about the patient, in addition to mechanisms like depicting the patient as untrustworthy or difficult. However, such studies have largely been limited to hand analysis of a small number of notes, limiting their generalizability to broader health data.

In this study, we used well-established NLP tools from computational social science and medical NLP to study evidence of racial bias in a large dataset of critical care clinical notes. Our primary hypothesis was that there are significant differences between critical care notes written for Black and White patients even after pairing based on patient age, gender, and primary diagnosis. We hypothesized that specific differences would be observed in both note style and note content, reflecting interpersonal and systemic elements of racism, respectively. In particular, we hypothesized that (1) notes for Black patients have a more negative emotional tone than notes for White patients, as well as higher levels of anxiety- and anger-related language; (2) notes for Black patients have more language related to social dominance, as measured by first personal plural pronouns and power-related language; and (3) there are significant differences in the mentions of opioid pain medication between Black and White patients, reflecting previously-observed systemic inequities in perception of pain and access to pain medication for Black patients.

## Materials and Methods

### Data

All data for this study comes from the Medical Information Mart for Intensive Care, version 3 (MIMIC-III), a public-use database of critical care admissions from Beth Israel Deaconess Medical Center in Boston, MA, between years 2001 to 2012. As the largest and most detailed public-use clinical database available, the iterative releases of the MIMIC dataset have been invaluable resources for clinical informatics research. In addition to its depth of structured data from critical care, including vital signs, lab reports, medications and procedures, etc., MIMIC includes a wealth of unstructured data in over two million free text clinical notes. These unstructured data are much more challenging to deidentify in securing clinical data for research purposes,^10^ making MIMIC central to research in medical NLP.

The most recent release of MIMIC to include free text notes, MIMIC-III, has been used to develop benchmark datasets for medical concept normalization,^11^ medical question answering,^12^ and medical natural language inference,^13^ as well as in developing clinical language models that are heavily used in current research.^14, 15^ Thus, if patterns of injustice— including implicit provider bias affecting patient interactions and care as well as structural injustice limiting access to high-quality care—are reflected in MIMIC data, then these patterns have the potential to be promulgated or exacerbated by NLP systems built on MIMIC’s foundation.^16, 17^

The representativeness of MIMIC clinical notes with respect to racial identity, and the racialized differences they reflect in the delivery of medical care, have not been investigated. This study presents an initial characterization of racialized differences in MIMIC documentation, and provides insights from our analyses into confounding factors and methodological challenges that may affect investigations into what clinical documentation reveals about the causes behind racial health disparities. By focusing on MIMIC, our analysis is reproducible by other researchers and shines an equity-focused light on a foundational resource for medical NLP research.

We used five tables from MIMIC-III. The *patients* table includes basic patient information such as gender (only available as binary male/female labels; gender assigned at birth was not explicitly recorded), date of birth, and date of death. The *admissions* table includes details related to each Intensive Care Unit (ICU) admission, such as admit time and discharge time, as well as patient demographics like ethnicity and insurance provider. We identified Black and White patients by extracting race from the *ethnicity* variable, which included both race and ethnicity (e.g., “WHITE – RUSSIAN” was mapped to “WHITE”). Primary ICD-9 codes for each admission were incorporated from the *diagnoses_icd* table, and the names of those diagnoses were taken from the *d_icd_diagnoses* table. Primary diagnoses were differentiated from secondary diagnoses by restricting to observations in the *diagnoses_icd* table where *seq_num* equaled 1. All clinical note information, including note text, is from the *noteevents* table. The full database consisted of 46,520 patients; 58,976 admissions; 2,083,180 notes, and 7,567 caregivers (e.g., physicians, nurses).

### Cohort Construction

We constructed racially-paired cohorts for our analysis. Because different stereotypes and biases exist for different races, we restricted to White or Black race only, allowing us to focus exclusively on pro-White, anti-Black racial bias. Additionally, MIMIC has few admissions for patients who are not Black or White (79.9% of admissions are Black or White), limiting our ability to conduct analyses with sufficient statistical power on other racial groups. We also restricted our sample to the two most common primary diagnoses by ICD-9 code, Unspecified Septicemia (ICD-9 code: 0389) and Coronary Atherosclerosis of Native Coronary Artery (ICD-9 code: 41401). We further stratified our sample into male and female cohorts to control for gender-related documentation differences. As our sample was not large enough to have grouped age cohorts, we restricted all cohorts to age 50 years and older to reduce age-related effects. This removed neonates from our sample and allowed us to focus on a relatively older population while still maintaining nearly 50 admissions in our smallest cohorts. We also dropped all notes marked as being made in error. The final sample consisted of 99,936 notes corresponding to 3,903 admissions of 3,748 patients. The notes were written by 1,230 unique caretakers, and our analysis included all available note categories (Table 1). This included all nursing and physician notes as well as notes from pharmacy, social work, rehab services, etc. Of this sample, 93.4% of patients were White and 66.4% were male. Sample stratification by race, gender, and diagnosis resulted in four pairs of cohorts, each pair differing by race (Table 2). We had 16 patients (2 Black, 14 White) who belonged to 2 cohorts, due to being admitted at separate times for different diagnoses. These patients represent 34 admissions (5 Black, 29 White) and 988 notes (51 Black, 937 White; <1% of all notes in the sample).

**Table 1.** Frequency of note categories within our sample.

| Note Type | Frequency |
| --- | --- |
| Nursing/other | 27,003 |
| Radiology | 22,062 |
| Nursing | 16,646 |
| ECG | 11,328 |
| Physician | 10,493 |
| Discharge Summary | 4,308 |
| Echo | 3,232 |
| Respiratory | 2,802 |
| General | 728 |
| Nutrition | 719 |
| Rehab Services | 372 |
| Social Work | 142 |
| Case Management | 92 |
| Consult | 5 |
| Pharmacy | 4 |

**Table 2.** Number of patients, admissions, and notes associated with each cohort.

| Race | Gender | Diagnosis | Patients | Admissions | Notes |
| --- | --- | --- | --- | --- | --- |
| White | Male | Septicemia | 664 | 712 | 27,777 |
| Black | Male | Septicemia | 70 | 81 | 2,579 |
| White | Male | Atherosclerosis | 1,717 | 1,728 | 31,256 |
| Black | Male | Atherosclerosis | 48 | 48 | 742 |
| White | Female | Septicemia | 617 | 659 | 21,741 |
| Black | Female | Septicemia | 86 | 103 | 3,920 |
| White | Female | Atherosclerosis | 516 | 524 | 11,119 |
| Black | Female | Atherosclerosis | 46 | 48 | 802 |
| Total |  |  | 3,748 | 3,903 | 99,936 |

### Vocabulary Analysis

We conducted an exploratory vocabulary analysis to get a sense of what types of words differed in relative frequency by race, and whether any of these words might reflect provider bias. We used SpaCy^18^ to tokenize the full note text for all available note categories, normalized each token to the lowercase version of its lemma, and restricted to alpha tokens only. We then calculated the frequency of each of these token lemmas by cohort and calculated the difference in relative frequency (normalized by number of admissions in the cohort) between matched cohorts. In browsing these frequencies, we identified notable differences in words that may reflect note writer bias towards the patient.

To conduct a more topic-based analysis beyond the level of individual words, we employed the Linguistic Inquiry and Word Count (LIWC) software, 2015 edition.^19^ LIWC is widely used in computational social science to analyze the frequency of various word categories, including emotion categories, social categories, and syntactic categories. LIWC includes 70 categories in total, and results are reported as the percentage of each note falling within a given category. LIWC allowed us to quantify the degree to which anger-related words are represented in a cohort’s notes, as well as other categories of words that may indicate bias such as emotional tone, negative emotion, and positive emotion. We also explored the themes of anxiety and social dominance, given their relevance in previous literature. We measured social dominance using the LIWC category of first person plural pronouns (such as in Hagiwara et al.^8^) and the *power* category, based on its description in the LIWC operator’s manual as “references relevant to status, dominance, social hierarchies.”^20^ Information on our selected LIWC categories are provided in Table 3.

**Table 3.** LIWC categories used for clinical note text analysis. The *emotional tone* category is a non-transparent variable calculated by aggregating over multiple categories, so it has no associated library of words.

| Category | Example Words | Words in Category |
| --- | --- | --- |
| Emotional Tone | N/A | N/A |
| Positive Emotion | love, nice, sweet | 620 |
| Negative Emotion | hurt, ugly, nasty | 744 |
| Anxiety | worried, fearful | 116 |
| Anger | hate, kill, annoyed | 230 |
| Power | superior, bully | 518 |
| 1st Per Plural Pron | we, us, our | 12 |

By applying LIWC software to the note text in our sample, we obtained the percentage of each note belonging to our categories of interest. The category of *emotional tone* is the exception, as that category is not reported as percentage of note, but rather a number from 0 to 100, with 0 being totally negative in tone, 50 being neutral, and 100 being totally positive. We then used the Mann-Whitney U test on these note-level observations with significance threshold *p<*0.05 to test whether each Black/White pair of cohorts differed in their percentage of note text in these categories. The Mann-Whitney U Test was chosen due to the non-normal distribution of the data, given that usually only a small amount of each note’s text belongs to a given LIWC category, if at all.

### Analysis of Opioid Mentions

To measure whether the relative mentions of pain medications significantly differed by patient race, we used the Apache cTAKES^21^ software (version 4.0.0) to identify the clinical concepts in each note and map them to the Unified Medical Language System^22^ (UMLS). cTAKES locates clinical concepts in a note and returns a UMLS Concept Unique Identifier (CUI) and a polarity value (negated terms have a polarity of -1). We created an *opioid* indicator that equaled 1 if the admission contained a note that mentioned any of the most common opioid analgesics used in the ICU, including morphine, fentanyl, hydromorphone, oxycodone, and methadone. This list was created with guidance from the clinical decision making reference, UpToDate,^23^ and opioids were identified in the data by tagging CUIs for which the preferred name contained the string *opioid, morphine, fentanyl, hydromorphone, remifentanil* (which had no matches), *oxycodone*, or *methadone*. We then ran a logistic regression model with robust standard errors (which account for any heteroskedasticity in the sample) to test whether race is significantly correlated with the likelihood of an admission containing an opioid mention in a note (Equation 1). Because negative opioid mentions in the notes were less likely, compared to positive mentions, to be reflections of opioids administered or prescribed, a second version of the regression was run. Here, the independent variable equaled 1 if the admission contained a note mentioning an opioid and the polarity equaled 1, unlike the original regression which had no polarity restriction.

**Equation 1**. Logistic regression of *opioid* on admission characteristics. The indicator *opioid* equaled 1 if the clinical note mentioned an opioid, 0 else; *black* equaled 1 if the patient was Black, 0 if White; *female* equaled 1 if the patient was female, 0 if male; *septicemia* equaled 1 if the primary diagnosis of the admission was septicemia, 0 if atherosclerosis. Model coefficients are represented by *β*_1_ to *β*_3_ and *β*_0_ is the intercept.

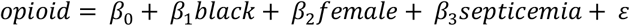

## Results

### Vocabulary Analysis

From our exploratory vocabulary analysis, we found there were many words with large frequency differences between racially paired cohorts. Some of these were indicative of health differences (e.g., *diabetes* and *renal* were much more common in notes for Black patients, reflecting the higher rates of diabetes and kidney disease in the Black population), and others seemed non-meaningful (e.g., *and* and *for* were more common in notes for White patients). Anger-related words such as *rude, belligerent, uncooperative*, and *aggressive* stood out as possible indicators of racial bias (Table 4).

**Table 4.** Vocabulary analysis results: examples of potential indicators of bias in male septicemia cohorts.

| Word | Race | Frequency | Cohort Admits | Freq/Cohort Admits |
| --- | --- | --- | --- | --- |
| Rude | White | 1 | 712 | 0.0014 |
|  | Black | 36 | 81 | 0.44 |
| Belligerent | White | 16 | 712 | 0.022 |
|  | Black | 36 | 81 | 0.44 |
| Uncooperative | White | 38 | 712 | 0.053 |
|  | Black | 25 | 81 | 0.31 |
| Aggressive | White | 1409 | 712 | 1.98 |
|  | Black | 244 | 81 | 3.01 |

In notes for male septicemia patients, the word *rude* appeared 36 times in 81 admissions for Black patients, but only once in 712 White admissions (a ∼300-fold difference). Among the same sample, *belligerent* was mentioned 36 times in 81 Black admissions, and 16 times in 712 White admissions (a ∼20-fold difference); *uncooperative* was mentioned 25 times in 81 Black admissions, and 38 times in 712 White admissions (a ∼6-fold difference); *aggressive* was mentioned 244 times in 81 Black admissions, and 1409 in 712 White admissions (a ∼1.5-fold difference).

To evaluate the validity of our vocabulary analysis, we conducted a manual review of all notes containing the word *rude*. This manual review provided several possible contributing factors for the use of the word *rude* in addition to possible note writer bias (Table 5). In particular, the copy-and-pasting of notes had a major influence, as the entirety of the differences seen in our exploratory vocabulary analysis were the result of a single sentence in the Black, male septicemia cohort getting copied across 36 notes. Furthermore, this same sentence accounted for all uses of *belligerent* and 36 of the 244 uses of *aggressive* in that cohort.

**Table 5.** Descriptions of the use of the word *rude* in clinical notes.

| Theme | Example |
| --- | --- |
| Possible negative attitude towards patient | "Patient has been totally appropriate tonight, only **rude** and stubborn. He wants things when he wants them, doesn't wait, gets [out of bed] without assistance even though he is told not to." |
| Description of non-patient | "One son esp, not following rules ie calling before entering unit, is loud, angry and **rude** to [name] and MD's" |
| Reporting what someone else said | "Floor nurse reported that pt. was very anxious and verbally **rude** due to annoyance with alarms and inability to get sleep." |
| Describing a disease with mental/emotional symptoms | "At times can be **rude** with nurses....Probably a combination of uremic/hepatic encephalopathy in the of sepsis." |
| Copy + paste | "Mental status: Pt was seen to be occasionally confused, saying odd things, and sometimes belligerent, making aggressive/**rude** comments to staff, and other times non-compliant, taking off BP cuff, pressing the pump buttons, not staying in bed, etc. Could be related to hepatic encephalopathy vs personality disorder." (repeated in 36 notes) |

### LIWC category analysis

We observed several types of statistically significant differences in the prevalence of LIWC categories between clinical notes for our paired cohorts, displayed in Table 6. (1) Notes for Black patients had more negative overall emotional tone than notes for White patients (male septicemia: p = 0.000, female septicemia: p = 0.000) and, separately, fewer positive emotion words (male septicemia: p = 0.000, female septicemia: p = 0.046, female atherosclerosis: p = 0.009) and more negative emotion words (male septicemia: p = 0.000, male atherosclerosis: p = 0.039, female septicemia: p = 0.001). (2) There were correlations between patient race and anxiety language (male septicemia: p = 0.000, male atherosclerosis: p = 0.009), and weak correlations between race and anger language (female septicemia: p = 0.017). However, these results were opposite of the expected direction, as all significant findings showed greater anxiety and anger in notes for White patients. (3) Notes for Black patients had higher levels of social dominance-related language, as measured by LIWC’s *power* category (male septicemia: p = 0.000, female septicemia: p = 0.000) and first person plural pronouns in the male septicemia cohorts (p = 0.000). However, the results for the female cohorts showed greater use of first person plural pronouns in notes for White patients (female septicemia: p = 0.013, female atherosclerosis: p = 0.026).

**Table 6.** Differences in cohort-level means for LIWC categories. The *B* columns represent the mean percentage of the given LIWC category in the relevant Black cohort’s notes, and the *W* columns represent the mean percentage of the given LIWC category in the relevant White cohort’s notes. The category of *emotional tone* is the exception, as that category is not reported as percentage of note, but rather a number from 0 to 100, with 0 being totally negative in tone, 50 being neutral, and 100 being totally positive. The *B-W* columns represent the value of the relevant *B* column minus the paired *W* column. Significance stars represent the p-values from Mann-Whitney U Tests. *** p<0.001, ** p<0.01, * p<0.05.

|  | Male Septicemia |  |  |  | Male Atherosclerosis |  |  |  | Female Septicemia |  |  |  | Female Atherosclerosis |  |  |  |
| --- | --- | --- | --- | --- | --- | --- | --- | --- | --- | --- | --- | --- | --- | --- | --- | --- |
|  | B | W | B-W | p | B | W | B-W | p | B | W | B-W | p | B | W | B-W | p |
| **Emotional Tone** | 28.305 | 30.282 | - | *** | 29.234 | 30.813 | - |  | 28.811 | 31.222 | - | *** | 28.816 | 30.356 | - |  |
| **Positive Emotion** | 1.486 | 1.556 | - | *** | 1.400 | 1.457 | - |  | 1.475 | 1.581 | - | * | 1.383 | 1.461 | - | ** |
| **Negative Emotion** | 1.721 | 1.553 | + | *** | 1.568 | 1.469 | + | * | 1.588 | 1.507 | + | ** | 1.477 | 1.535 | - |  |
| **Anxiety** | 0.209 | 0.235 | - | *** | 0.168 | 0.191 | - | ** | 0.214 | 0.245 | - |  | 0.187 | 0.204 | - |  |
| **Anger** | 0.074 | 0.067 | + |  | 0.046 | 0.055 | - |  | 0.043 | 0.063 | - | * | 0.047 | 0.044 | + |  |
| **Power** | 2.269 | 2.096 | + | *** | 2.214 | 2.136 | + |  | 2.233 | 2.139 | + | *** | 2.044 | 2.169 | - |  |
| **1st Per Plural Pron** | 0.048 | 0.030 | + | *** | 0.006 | 0.010 | - |  | 0.032 | 0.035 | - | * | 0.008 | 0.013 | - | * |

There was a great deal of variance among the individual clinical notes in terms of LIWC values. As an example of this variance, the distribution of LIWC categories for three notes in the same admission (*hadm_id* = 100009; from the White, male atherosclerosis cohort) is shown in Figure 1. No two example notes share the same set of observed categories, and the frequency of each category varies widely.

**Figure 1.**
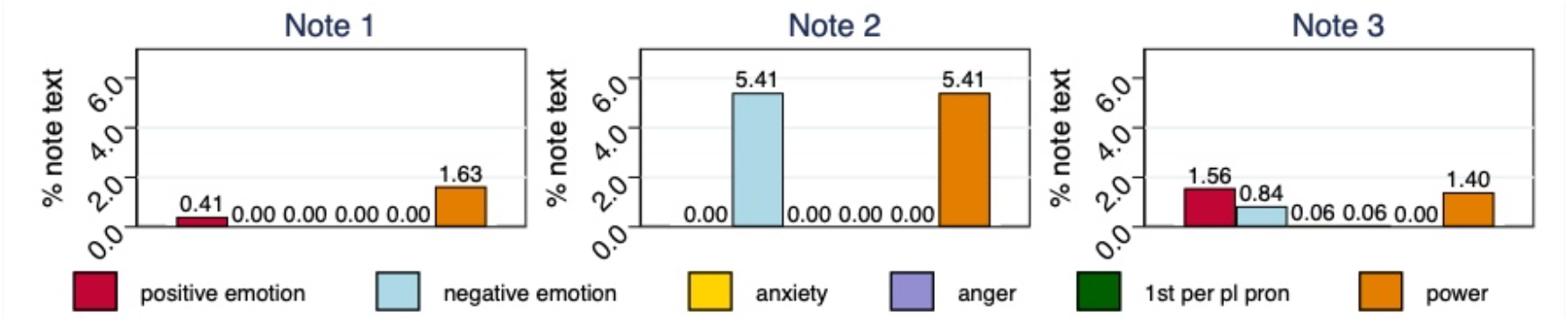
The distribution of LIWC categories in three notes from a single admission (*hadm_id* = 100009; from the White, male atherosclerosis cohort). The *emotional tone* category was excluded because it is not reported as a percentage of note text.

Each category is only observed in a subset of the notes, although all of our selected categories were observed in each cohort. Some categories, like *negative emotion*, were found in nearly all notes, while others, such as *anger*, were observed in less than 20% of notes (Figure 2). To understand the trends in LIWC values when they are observed, we graphed the nonzero values for each LIWC category by cohort (results for *negative emotion* and *anger* shown in Figure 3; other categories exhibited similar patterns) and measured significant differences between cohorts using Mann-Whitney U tests. Means and medians remained similar between the paired cohorts, but the spread of the values varied considerably, particularly in the first and fourth quartiles. Thus, small sets of notes with unusually high or low category frequencies were the primary factors distinguishing the cohorts, rather than systematic trends.

**Figure 2.**
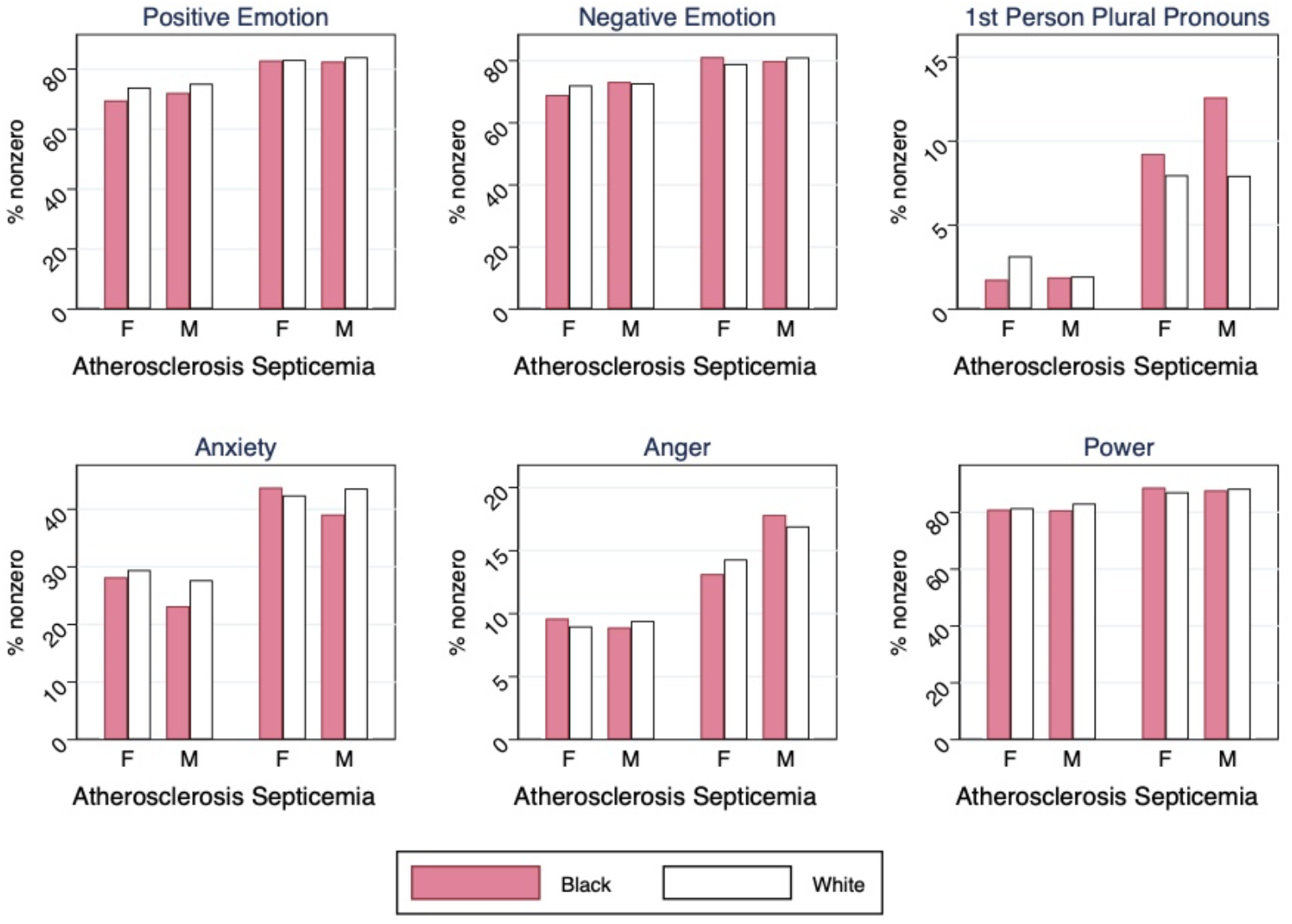
The percentage of notes in each cohort for which a nonzero proportion of the words were tagged within each LIWC category.

**Figure 3.**
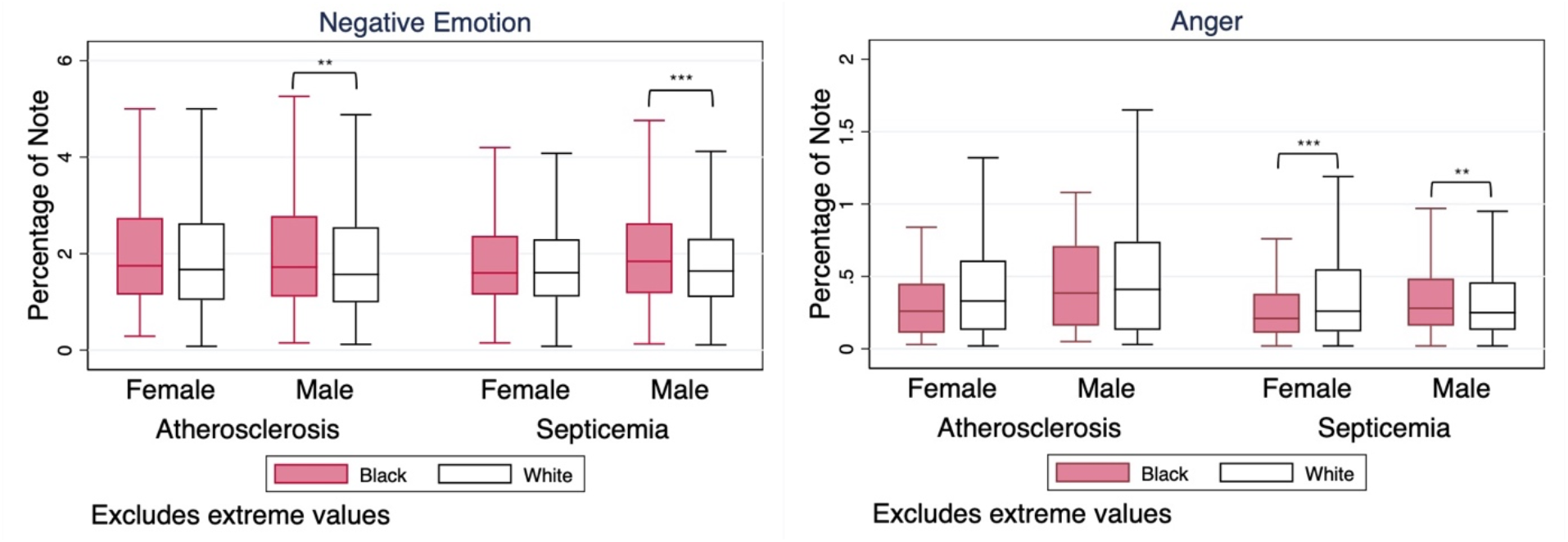
Distribution of LIWC categories *negative emotion* and *anger*, by cohort. Box-and-whisker plots were created using nonzero values only and extreme values (outside 1.5 times the interquartile range) are excluded. Significance stars are p-values of Mann-Whitney U Tests on nonzero observations. *** p<0.001, ** p<0.01, * p<0.05.

### Analysis of Opioid Mentions

Fourteen CUIs representing opioids were identified in the clinical notes. The most mentioned were fentanyl (N = 13,648), morphine (N = 10,925), and oxycodone (N = 3,996). Nearly all mentions (99.8%) had a positive polarity. The full list of identified CUIs and their frequencies are listed in Table 7.

**Table 7.**
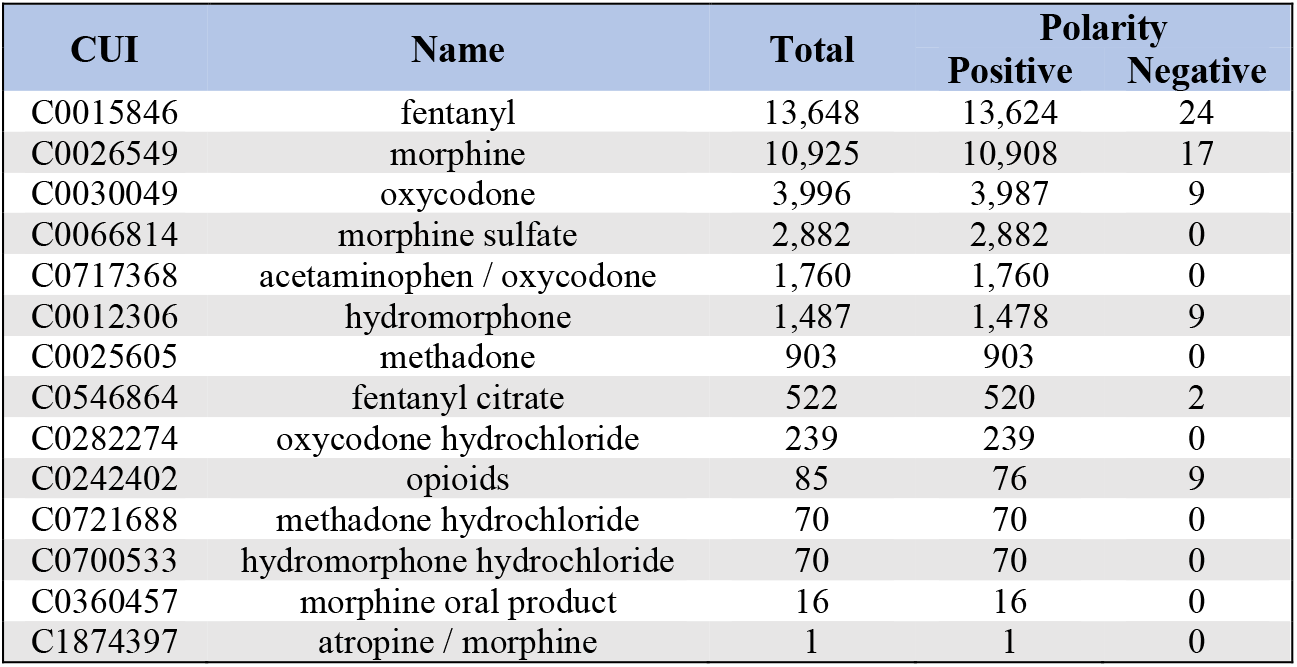
Opioid-related CUIs identified in the clinical note text and their frequencies.

| CUI | Name | Total | Polarity |  |
| --- | --- | --- | --- | --- |
|  |  |  | Positive | Negative |
| C0015846 | fentanyl | 13,648 | 13,624 | 24 |
| C0026549 | morphine | 10,925 | 10,908 | 17 |
| C0030049 | oxycodone | 3,996 | 3,987 | 9 |
| C0066814 | morphine sulfate | 2,882 | 2,882 | 0 |
| C0717368 | acetaminophen / oxycodone | 1,760 | 1,760 | 0 |
| C0012306 | hydromorphone | 1,487 | 1,478 | 9 |
| C0025605 | methadone | 903 | 903 | 0 |
| C0546864 | fentanyl citrate | 522 | 520 | 2 |
| C0282274 | oxycodone hydrochloride | 239 | 239 | 0 |
| C0242402 | opioids | 85 | 76 | 9 |
| C0721688 | methadone hydrochloride | 70 | 70 | 0 |
| C0700533 | hydromorphone hydrochloride | 70 | 70 | 0 |
| C0360457 | morphine oral product | 16 | 16 | 0 |
| C1874397 | atropine / morphine | 1 | 1 | 0 |

The results of the logistic regression (Equation 1) indicated that notes for Black patients were less likely to mention opioids relative to notes for White patients (odds ratio = 0.685, p = 0.003). The results of the second regression, in which the dependent variable equaled 1 if an opioid was mentioned <u>and</u> the polarity was positive, were nearly identical to the original regression (odd ratio = 0.687, p = 0.004). Opioids (as listed in Table 7) were commonly mentioned, occurring in over 50% of the notes in each cohort (Figure 4). Opioid mentions were more common for White patients in all cohorts except female atherosclerosis.

**Figure 4.**
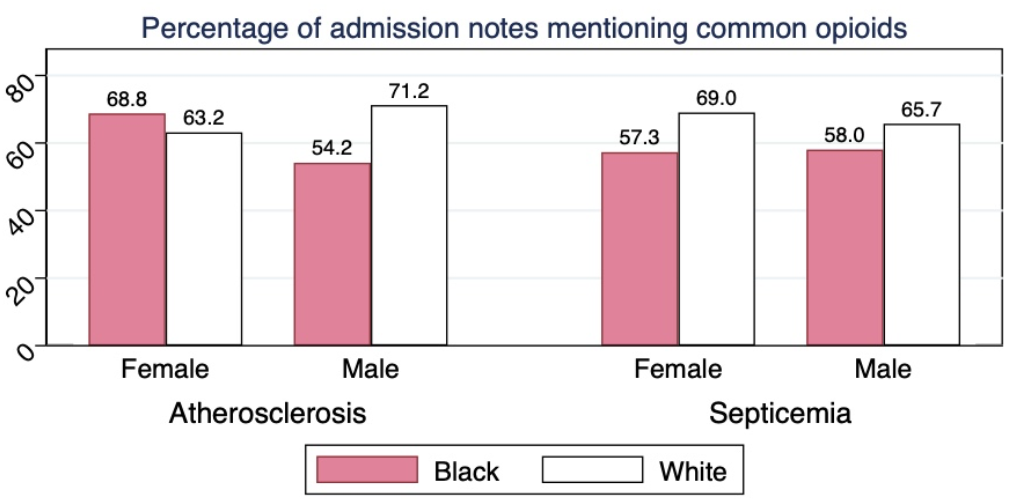
Percentage of admission notes mentioning common ICU-administered opioids, by cohort.

## Discussion

### Vocabulary and LIWC Analyses

The results of our analyses show significant differences in language used for Black and White patients in clinical notes. Notes for Black patients appeared to have a more negative emotional tone than notes for White patients, as reflected by differences in overall emotional tone as well as differences in positive and negative emotion. This could be a reflection of clinician bias, given Park et al.’s finding that negative emotional language is a form of stigmatizing language in clinical notes.^9^

We also observed higher levels of social dominance-related language in notes for Black patients. Like Hagiwara et al.,^8^ we did find significant differences in first person plural pronoun use between Black and White cohorts. However, the direction was not always consistent across cohort pairs or with previous findings. We found that for female patients with a septicemia or atherosclerosis diagnosis, there was greater use of first person plural pronouns in notes for White patients compared to those for Black patients. For male patients, we found significantly fewer mentions of such pronouns for White septicemia patients and no significant difference for male atherosclerosis patients.

Our other measure of social dominance, LIWC’s *power* category, also represented a higher percentage of each note for Black patients, compared to White. Results for anxiety- and anger-related language were opposite of what we expected based on previous work and our vocabulary analysis, as all significant findings showed lower levels of anxiety or anger in notes for Black patients compared to White. However, the results for anger were only statistically significant for female septicemia patients.

### Validity of LIWC

To dig deeper into these findings and assess the degree to which they indicate potential provider bias, we reviewed the top words tagged in each LIWC category. **We found that for some categories, LIWC was unable to differentiate between clinical language and words that may reflect bias**. For example, in the male septicemia cohorts, the top words in the *negative emotion* category included *pain, failure, low, lower*, and *shock*. These words are typically used in the clinical context to describe patient health status, rather than reflections of provider attitude. Similarly, top words in the *power* category included *failure, status, up, low*, and *doctor*—each of which is again in common clinical usage without indications of social dominance expected in non-healthcare contexts. Specific categories, such as anger, seemed to be more accurate at capturing provider attitude, but more context is required to determine whether anger language is present in the note due to provider attitude towards the patient, or due to some other reason, such as a description of a patient’s family member or visitor. While a word frequency analysis necessarily elides contextual details, an LIWC-like tool tailored for clinical notes would nonetheless be a valuable technology for researchers studying implicit bias in clinical notes. This would require adapting categorical dictionaries to distinguish between words likely related to health, such as *pain*, and words more clearly related to affect, such as *belligerent*.

### Analysis of Opioid Mentions

Beyond stylistic findings, we also found an information content difference in that notes for Black patients are significantly less likely to mention an opioid. This could reflect racial bias, as it is an outcome consistent with the idea that health professionals underestimate and undertreat the pain Black patients experience. There are several possible explanations for why providers underestimate and undertreat Black patients’ pain, including incorrect provider beliefs about Black patients’ nerve endings and skin,^6^ or suspicion that Black patients are more likely to abuse opioids than White patients.^24, 25^ It is also possible that these results do not indicate bias, but instead represent racial trends in opioid abuse, since during this time period the opioid crisis disproportionately affected White Americans.^26^ Both of these factors are likely to be intertwined in practice. In order to gain more nuance in this analysis, including distinguishing between opioid mentions in the patient history and ICU administration of opioids, these results could be checked against the medication administration data in MIMIC-III.

### Implications

Overall, we find that as a method of investigating implicit bias in healthcare, applying computational analysis to clinical notes allows for faster analysis and the utilization of much larger datasets compared to hand analysis, but introduces additional challenges in accounting for the context and pragmatic understanding behind quantitatively observed differences. Developing more nuanced methods for computational analysis will be key to achieving the potential of computational techniques to gain insight into data of interest. For example, as computational techniques can be straightforwardly applied to any dataset without resource-intensive data curation, they can be used to evaluate and gain insight into implicit bias levels and mechanisms for teams, departments, or entire institutions. This could inform targeted interventions to combat implicit bias at multiple levels of health professions training and practice. For example, if there are racialized differences in the use of negative emotional language in clinical notes, medical trainees may be taught about this difference, which could act as awareness intervention^27, 28^ that reduces bias in note writing. This would likely have positive downstream effects, as a reduction in biased notes would reduce the probability that a health professional would read a biased note and perceive a patient differently.

## Limitations

This study had several limitations which can inform future work on developing computational methods for analyzing evidence of bias at scale.

### Sample Limitations

The patient sample in MIMIC is strongly skewed White, and the dataset represents one well-resourced medical center in a major city in the Northeastern U.S. Additionally, all patients were critical care patients, who are sicker and likely more socioeconomically vulnerable than the overall population. Furthermore, patients in the ICU may be completely incapacitated and have limited interaction with providers. This may have a significant influence on provider attitude, by limiting interactions in which providers can form an opinion on the patient; conversely, this lack of conversation may make clinicians more likely to stereotype patients based on observable characteristics like skin color. We also limited our analysis to two primary diagnoses, representing a relatively small subset of the overall patient population. Diagnosis is likely to affect how clinicians interact with patients, as some diagnoses are more incapacitating than others (e.g., septicemia patients are usually far sicker than atherosclerosis patients), and some diagnoses have more behavioral manifestations than others. The racial skew of the data also limited the power of our analysis. A larger share of Black patients in the dataset would allow for further cohort stratification, controlling for factors like insurance provider or creating multiple age brackets. Additionally, the small size of the Black cohorts compared to the White cohorts reduced both statistical power and the diversity of data available to draw on. We tended to find more significant results in the male and female septicemia cohorts compared to the atherosclerosis cohorts, the latter of which had notably smaller Black populations both by absolute admission counts and relative to their White populations. Several of these limitations are inherent in the MIMIC dataset itself, highlighting important considerations of under-representation in NLP and other informatics work based on MIMIC data. Future research on characterizing—and potentially mitigating—implicit bias in clinical documentation will thus best be served by sampling datasets with explicit criteria for diverse representation of patient demographics, including race, age, and gender identity. In addition, techniques such as propensity score matching (in cohort construction) and structured equation modeling (in data analysis) can help to reduce the influence of confounding variables.

### Missing Note Writer Demographics

Information on note writer demographics can be important predictors for levels of implicit bias^29^ and would also be valuable to incorporate into comparative analyses. However, this information is included unreliably or not at all in MIMIC-III. Future study could apply similar methods to ours to identifiable data within a healthcare system, which would allow for the controlling of clinician characteristics.

### Copy-and-Pasting of Note Text

As highlighted by our manual review, the copy-and-pasting of note text across multiple notes has the potential to substantially distort analyses relying on word counts. A recent study by Rule et al^30^ describes a potential method for identifying these occurrences which can be employed in future work. Additionally, the diversity of note lengths and content types in MIMIC-III notes—ranging from a few dozen words noting an encounter to extensive documentation of history and physical findings—affects LIWC values and may overweight some notes. Various strategies may be employed in future work for reducing the impact of these factors, including length-sensitive weighting and focused analyses of specific note types. For example, notes focused on objective measures and tests, such as radiology notes, may be less likely to reflect bias given the limited interaction between the note author and the patient.

## Conclusion

This study investigated the use of computational methods to study racial bias in a large, heterogenous dataset of clinical note text. Computational analysis identified significant differences in note style and content between Black and White patients, including that notes for Black patients had more negative emotional tone, greater use of social dominance language, and fewer mentions of opioid medications. We identified multiple potential factors contributing to these differences in addition to implicit bias, including mis-categorization of healthcare words as emotional in tone. Our findings do not suggest that the impact of implicit bias in healthcare is overestimated—rather, they illuminate the complexity and importance of effective measurement and detailed analysis of evidence of bias in healthcare practice. Our study showed that computational text analysis methods have significant potential for characterizing racial differences in clinical documentation, and identified key design considerations for future research into the mechanisms of racial disparities in health documentation.

## Data Availability

All data produced in the present study are available upon reasonable request to the authors.

https://physionet.org/content/mimiciii/1.4/

## Acknowledgements

This research was performed during the first author’s participation in the Short-Term Trainee Program of the University of Pittsburgh Biomedical Informatics Training Program. Both authors were supported by the National Library of Medicine of the National Institutes of Health under award number T15LM007059.

## References

1. FitzGerald C, Hurst S. Implicit bias in healthcare professionals: a systematic review. BMC Med Ethics. 2017;18(1):19.

2. Green AR, Carney DR, Pallin DJ, Ngo LH, Raymond KL, Iezzoni LI, et al. Implicit bias among physicians and its prediction of thrombolysis decisions for black and white patients. J Gen Intern Med. 2007;22(9):1231–8.

3. Sabin JA, Greenwald AG. The influence of implicit bias on treatment recommendations for 4 common pediatric conditions: pain, urinary tract infection, attention deficit hyperactivity disorder, and asthma. Am J Public Health. 2012;102(5):988–95.

4. Heins JK, Heins A, Grammas M, Costello M, Huang K, Mishra S. Disparities in analgesia and opioid prescribing practices for patients with musculoskeletal pain in the emergency department. J Emerg Nurs. 2006;32(3):219–24.

5. Stepanikova I. Racial-ethnic biases, time pressure, and medical decisions. J Health Soc Behav. 2012;53(3):329–43.

6. Hoffman KM, Trawalter S, Axt JR, Oliver MN. Racial bias in pain assessment and treatment recommendations, and false beliefs about biological differences between blacks and whites. Proc Natl Acad Sci U S A. 2016;113(16):4296–301.

7. Beach MC, Saha S, Park J, Taylor J, Drew P, Plank E, et al. Testimonial Injustice: Linguistic Bias in the Medical Records of Black Patients and Women. J Gen Intern Med. 2021;36(6):1708–14.

8. Hagiwara N, Slatcher RB, Eggly S, Penner LA. Physician Racial Bias and Word Use during Racially Discordant Medical Interactions. Health Commun. 2017;32(4):401–8.

9. Park J, Saha S, Chee B, Taylor J, Beach MC. Physician Use of Stigmatizing Language in Patient Medical Records. JAMA Netw Open. 2021;4(7):e2117052.

10. Yang X, Lyu T, Li Q, Lee CY, Bian J, Hogan WR, et al. A study of deep learning methods for de-identification of clinical notes in cross-institute settings. BMC Med Inform Decis Mak. 2019;19(Suppl 5):232.

11. Denecke K. Sublanguage analysis of medical weblogs. Stud Health Technol Inform. 2014;205:565–9.

12. Roberts K, Simpson MS, Voorhees EM, Hersh WR, editors. Overview of the TREC 2015 Clinical Decision Support Track. TREC; 2015.

13. Romanov A, Shivade C. Lessons from natural language inference in the clinical domain. arXiv preprint 180806752. 2018.

14. Alsentzer E, Murphy JR, Boag W, Weng W-H, Jin D, Naumann T, et al. Publicly available clinical BERT embeddings. arXiv preprint 190403323. 2019.

15. Peng Y, Yan S, Lu Z. Transfer learning in biomedical natural language processing: an evaluation of BERT and ELMO on ten benchmarking datasets. arXiv. arXiv preprint 190605474. 2019.

16. Obermeyer Z, Powers B, Vogeli C, Mullainathan S. Dissecting racial bias in an algorithm used to manage the health of populations. Science. 2019;366(6464):447–53.

17. Bender EM, Gebru T, McMillan-Major A, Shmitchell S, editors. On the Dangers of Stochastic Parrots: Can Language Models Be Too Big? Proceedings of the 2021 ACM Conference on Fairness, Accountability, and Transparency; 2021.

18. Honnibal M, Montani I. spaCy 2: Natural language understanding with Bloom embeddings, convolutional neural networks and incremental parsing. To appear. 2017;7(1):411–20.

19. Pennebaker JW, Boyd RL, Jordan K, Blackburn K. The development and psychometric properties of LIWC2015. 2015.

20. Pennebaker JW, Booth RJ, Boyd RL, Francis ME. Linguistic Inquiry and Word Count: LIWC2015 Operator’s Manual. Austin, TX; 2015.

21. Savova GK, Masanz JJ, Ogren PV, Zheng J, Sohn S, Kipper-Schuler KC, et al. Mayo clinical Text Analysis and Knowledge Extraction System (cTAKES): architecture, component evaluation and applications. J Am Med Inform Assoc. 2010;17(5):507–13.

22. Bodenreider O. The Unified Medical Language System (UMLS): integrating biomedical terminology. Nucleic Acids Res. 2004;32(Database issue):D267–70.

23. Tietze KJ, Fuchs B. Sedative-analgesic medications in critically ill adults: properties, dosage regimens, and adverse effects. Waltham, MA: UpToDate Accessed on February. 2016;25.

24. Becker WC, Starrels JL, Heo M, Li X, Weiner MG, Turner BJ. Racial differences in primary care opioid risk reduction strategies. Ann Fam Med. 2011;9(3):219–25.

25. Hirsh AT, Anastas TM, Miller MM, Quinn PD, Kroenke K. Patient race and opioid misuse history influence provider risk perceptions for future opioid-related problems. Am Psychol. 2020;75(6):784–95.

26. Becker WC, Sullivan LE, Tetrault JM, Desai RA, Fiellin DA. Non-medical use, abuse and dependence on prescription opioids among U.S. adults: psychiatric, medical and substance use correlates. Drug Alcohol Depend. 2008;94(1-3):38–47.

27. Sherman MD, Ricco J, Nelson SC, Nezhad SJ, Prasad S. Implicit Bias Training in a Residency Program: Aiming for Enduring Effects. Fam Med. 2019;51(8):677–81.

28. Zeidan AJ, Khatri UG, Aysola J, Shofer FS, Mamtani M, Scott KR, et al. Implicit Bias Education and Emergency Medicine Training: Step One? Awareness. AEM Educ Train. 2019;3(1):81–5.

29. Sabin J, Nosek BA, Greenwald A, Rivara FP. Physicians’ implicit and explicit attitudes about race by MD race, ethnicity, and gender. J Health Care Poor Underserved. 2009;20(3):896–913.

30. Rule A, Bedrick S, Chiang MF, Hribar MR. Length and Redundancy of Outpatient Progress Notes Across a Decade at an Academic Medical Center. JAMA Netw Open. 2021;4(7):e2115334.

